# FIRST DETECTION OF SARS-COV-2 IN UNTREATED WASTEWATERS IN ITALY

**DOI:** 10.1101/2020.04.25.20079830

**Authors:** Giuseppina La Rosa, Marcello Iaconelli, Pamela Mancini, Giusy Bonanno Ferraro, Carolina Veneri, Lucia Bonadonna, Luca Lucentini, Elisabetta Suffredini

**Author notes:** Corresponding author: Giuseppina La Rosa, Istituto Superiore di Sanità, Department of Environment and Health, Viale Regina Elena 299, 00161, Rome, Italy.

## Abstract

Several studies have demonstrated the advantages of environmental surveillance through the monitoring of sewer systems for the assessment of viruses circulating in a given community (wastewater-based epidemiology, WBE).

During the COVID-19 public health emergency, many reports have described the presence of SARS-CoV-2 RNA in stools from COVID-19 patients, and a few studies reported the occurrence of SARS-CoV-2 in wastewaters worldwide. Italy is among the world’s worst-affected countries in the COVID-19 pandemic, but so far there are no studies assessing the presence of SARS-CoV-2 in Italian wastewaters. To this aim, twelve influent sewage samples, collected between February and April 2020 from Wastewater Treatment Plants in Milan and Rome, were tested adapting, for concentration, the standard WHO procedure for Poliovirus surveillance. Molecular analysis was undertaken with three nested protocols, including a newly designed SARS-CoV-2 specific primer set.

SARS-CoV-2 RNA detection occurred in volumes of 250 mL of wastewaters collected in both areas at high (Milan) and low (Rome) epidemic circulation, according to clinical data. Overall, 6 out of 12 samples were positive. One of the positive results was obtained in a Milan wastewater sample collected a few days after the first notified Italian case of autochthonous SARS-CoV-2.

The study shows that WBE has the potential to be applied to SARS-CoV-2 as a sensitive tool to study spatial and temporal trends of virus circulation in the population.

## INTRODUCTION

Severe Acute Respiratory Syndrome Coronavirus 2 (SARS-CoV-2) is responsible for the coronavirus disease COVID-19, a public health emergency worldwide. On March 11 2020, the World Health Organization declared COVID-19 a pandemic. Italy is among the world's most affected countries in the COVID-19 pandemic. Indeed, after entering Italy, COVID-19 has been spreading fast. As of April 20^th^ 2020, the total number of cases reported by the authorities reached 181,228, with 108,237 active cases (Dipartimento della Protezione Civile, Bulletin 20.04.2020), mainly located in Northern Italy (Lombardy, and its neighbouring regions of Emilia-Romagna and Piedmont).

Presymptomatic and paucisymptomatic carriers, mostly undetected in clinical and laboratory surveillance systems, contribute to the spread of the disease (Bay et al., 2020; Nicastri et al., 2020; Rothe et al., 2020; WHO, 2020) and hamper the efforts made to assess the extent of SARS-CoV-2 circulation in the population and to control efficiently virus transmission. Analytical regular investigation of wastewaters provides valuable information to measure viral circulation in the population as Wastewater Treatment Plants (WWTPs), collecting and concentrating human excreta, are useful sampling points receiving discharges from the entire community.

Environmental microbiologists have studied pathogens in sewage for decades (La Rosa & Muscillo, 2012; Sinclair *et al*., 2008). The screening of wastewater, as a public health surveillance tool, defined as wastewater-based epidemiology (WBE), is currently well recognized (Xagoraraki & O'Brien, 2020). In the recent years, scientists have applied WBE to a wide range of waterborne, foodborne and fecal-oral viruses, which infected individuals usually excrete in high concentration with faeces (Katayama et al., 2008; Iaconelli et al., 2017; Bisseaux et al., 2018). However, the concept of WBE can also be applied to viruses beyond those commonly associated with the faecal-oral route (i.e. enteric viruses), since viral shedding may involve different body fluids ultimately discharged into urban sewage.

Some studies have reported the presence of virus RNA in the stools of COVID-19 patients in percentages ranging from 16.5% to 100% at a concentration up to 6.8 log_10_ genome copies/g of stool (Chen et al., 2000; Lo et al., 2000; Han et al., 2000; Lescure et al., 2000). Furthermore, preliminary studies have reported the detection of SARS-CoV-2 RNA in wastewater in The Netherlands (Medema *et al*., 2020), France (Wurtzer et al., 2020), USA (Wu et al., 2020), and Australia (Ashmed et al., 2020). To date, no study has yet provided insights into the presence of SARS-CoV-2 in wastewaters in Italy.

Herein we report the results of the screening for SARS-CoV-2 presence in sewage samples collected between the end of February and the beginning of April 2020 from WWTPs in Milan (Northern Italy) and Rome (Central Italy).

## MATERIAL AND METHODS

Twelve raw sewage samples were collected between the 3^rd^ of February and the 2^nd^ of April 2020 from three WWTPs, located in Milan (two distinct plants, reported as A and B) and in Rome (one plant receiving two different pipelines, C1 and C2, from different districts of the town), respectively. The samples (24-hour composites) were collected from the WWTP influent, immediately stored at -20 °C, and dispatched frozen to the National Institute of Health for analysis. Before viral concentration, samples underwent a 30 min treatment at 57 °C to increase the safety of the analytical protocol for the laboratory personnel and environment, as Pastorino et al. (2020) reported these conditions to reduce the virulence of the virus by over 5 log.

Sample concentration took place using a two-phase (PEG-dextran method) separation as detailed in the 2003 WHO Guidelines for Environmental Surveillance of Poliovirus protocol (WHO, 2003), with modifications to adapt the protocol to enveloped viruses. In brief, the wastewater sample (250 mL) was centrifuged to pellet the wastewater solids, retaining the pellet for further processing. The clarified wastewater was mixed with dextran and polyethylene glycol (PEG), and the mixture was left to stand overnight at 4 °C in a separation funnel. The bottom layer and the interphase were then collected drop-wise, and this concentrate was added to the pellet from the initial centrifugation. The chloroform treatment that the WHO protocol envisages at this stage was omitted; this preserved the integrity of the enveloped viruses object of this study. The viral nucleic acids extraction foresaw the use of the NucliSENS miniMAG semi-automated extraction system with magnetic silica carried out following manufacturer’s instructions (bioMerieux, Marcy l’Etoile, France) with however slight modifications. The lysis phase was prolonged to 20 minutes, and brief centrifugation (2000 × *g*, 1 min) was used to pellet the sediment; subsequently, magnetic silica beads were added to the cleaned supernatant. Before molecular tests, the extracted nucleic acids were further purified by PCR inhibitors using the OneStep PCR Inhibitor Removal Kit (Zymo Research, CA, USA).

In the absence of a standardized method for SARS-CoV-2 detection in environmental samples, RNAs were tested for the presence of SARS-CoV-2 using three different PCR assays (Table 1 and Figure 1):

a. a broad range of PCR for Coronavirus detection targeting the ORF1ab (Ar Gouilh et al., 2018). Primers were previously designed targeting a highly conserved region (nsp12) among all Coronavirinae sequences to detect a broad range of coronaviruses by a semi-nested PCR producing a fragment of 218 bp.
b. a newly designed primer set specific for SARS-CoV-2. Novel nested primers, amplifying a 332 bp fragment of ORF1ab, were designed using prime3 software (http://primer3.ut.ee/). For the assays a) and b) first-strand cDNA was synthesized using Super Script IV Reverse Transcriptase (ThermoFisher Scientific) with the reverse primer. PCR reaction was performed using 2.5 μl of cDNA in a final volume of 25 μl (Kit Platinum^™^ SuperFi^™^ Green PCR Master Mix, Thermo), using 1 µl of primers (10 µM). The PCR conditions were as follows: 98 °C for 30 sec; 35 cycles of 98 °C for 10 sec, 50 °C and 54 °C for 10 sec for assay a) and b) respectively, and 72 °C for 30 sec; final extension 72 °C for 5 min. After the first round PCR, nested PCR was performed using 2 µl of first PCR product and under the same reaction composition and thermal profile conditions. A synthetic DNA (Biofab Research) including the PCR target region, was used to set up PCR conditions before experiments with study samples, but was not amplified along with samples to avoid risks of PCR contamination. Molecular grade water was used as the negative control.
c. a published nested RT-PCR for SARS-CoV-2 targeting the spike region (Nao et al., 2020). cDNA was synthesized from 5 μl of sample RNA, using SuperScript III Reverse Transcriptase (ThermoFisher Scientific), 0.5 μM of the reverse primer (WuhanCoV-spk2-r, Table 1) and a 50 min reaction at 50 °C (20 μl final volume). First PCR reaction was performed by adding the reaction mix (Dream Taq polymerase and buffer from ThermoFisher Scientific, 0.4 μM of primers WuhanCoV-spk2-r and WuhanCoV-spk1-f) directly to the whole volume of synthesized cDNA. The used PCR conditions were as follows: 95°C for 1 min; 35 cycles of 95 °C for 30 sec, 56 °C for 30 sec, and 72 °C for 40 sec; final extension 72 °C for 5 min. Nested PCR (primers NIID_WH-1_F24381 and NIID_WH-1_R24873) was performed in a total volume of 50 µl using 5 µl of first PCR product, with the same conditions applied for the first PCR and 45 cycles.

**Table 1:**
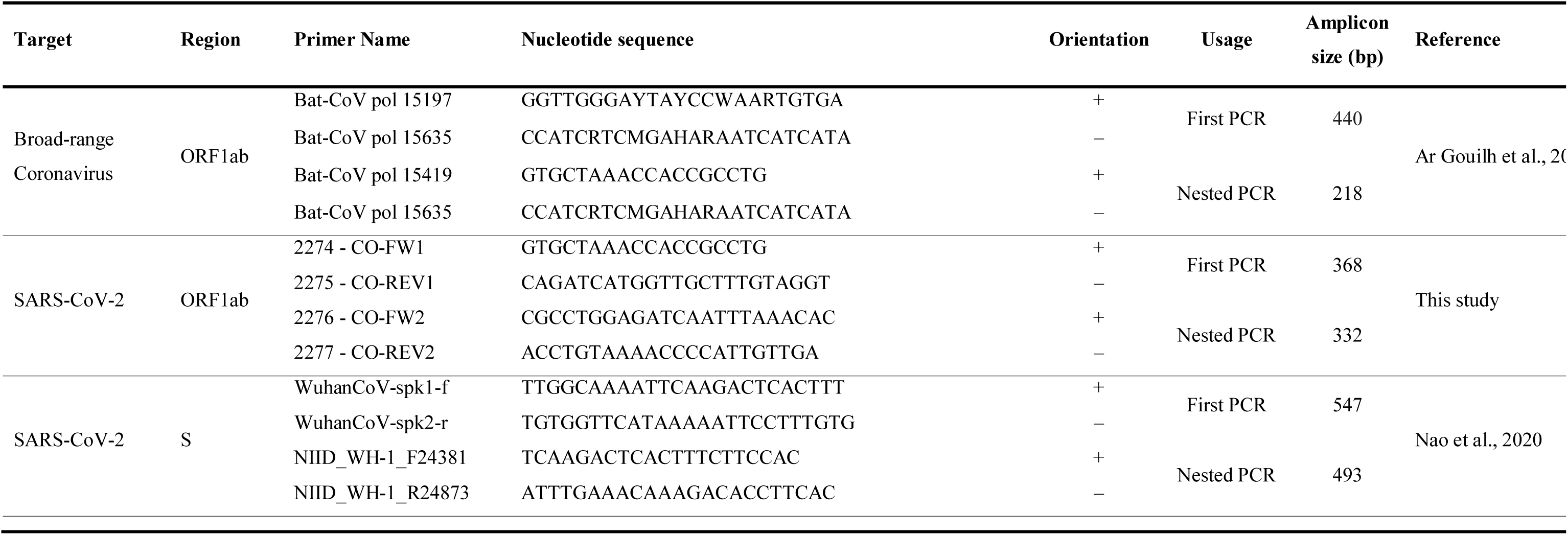
Primers and amplification protocols used in the study

**Fig. 1:**
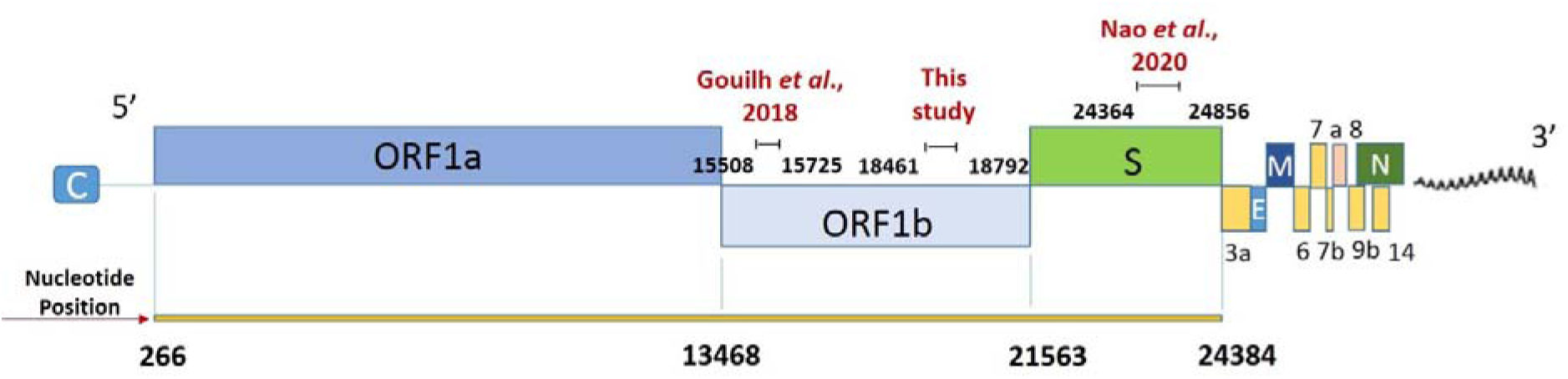
Position on SARS-CoV-2 genome of the primers used in the study. Modified from Viralzone (https://viralzone.expasy.org/9076)

All positive samples were retested for confirmation of results. The PCR products were revealed by electrophoresis on 2% agarose gels and were purified using a Montage PCRm96 Microwell Filter Plate (Millipore, Billerica, MA, USA) and then direct sequenced on both strands (BioFab Research, Rome, Italy). Sequences were identified in terms of the closest homology sequence using BLAST https://blast.ncbi.nlm.nih.gov/Blast.cgi. All Italian SARS-CoV-2 genome sequences available at the time of analysis were retrieved from Gisaid (https://www.gisaid.org/) for comparison with study sequences, using the MEGA X software (Kumar *et al*., 2018).

Sequences were submitted to NCBI GenBank with the accession numbers: [GenBank submission ID 2337138 and 2337168].

## RESULTS AND DISCUSSION

The 50% (6/12) of the wastewater samples showed positive results for SARS-CoV-2 RNA (Table 2). Both the published and newly designed SARS-CoV-2 specific primer sets detected bands of the expected size and were confirmed by the sequencing. In contrast, only aspecific products were detected with a broad range assay for coronavirus. Upon comparison of broad range primers with SARS-CoV-2 genome, we noted that they showed only 77.1 to 91.3 % nt identity, which explain why these were not able to amplify the novel coronavirus.

**Table 2:** Results of SARS-CoV-2 detection in the study period

| City and WWTP | Date of sampling |  |  |  |  |  |  |  |
| --- | --- | --- | --- | --- | --- | --- | --- | --- |
|  | 03 Feb | 19 Feb | 23 Feb | 24 Feb | 26 Feb | 28 Feb | 31 Mar | 02 Apr |
| Milan – plant A |  | × |  | ○<sup>a</sup> | × | × |  |  |
| Milan – plant B | × |  | × |  | × | ○● |  |  |
| Rome – plant C1 |  |  |  |  |  |  | ○<sup>a</sup> | ○ |
| Rome – plant C2 |  |  |  |  |  |  | ○ | ○● |

× SARS-CoV-2 not detected; ○ SARS-CoV-2 detected (ORF1ab); ● SARS-CoV-2 detected (spike)
<sup>a</sup> weak positive

SARS-CoV-2 RNA was first detected with a weak amplification signal in an influent sample from Milan WWTPs (Lombardy, North Italy) collected on February 24^th^ from the plant A with the ORF1ab assay. A positive result for SARS-CoV-2 was also detected this time with a strong amplification signal, on February 28^th^ from the plant B, with both the ORF1ab and the Spike regions. In the influent samples taken from the WWTP in Rome (Latium, Central Italy), SARS-CoV-2 was detected in both the sampling dates (31^st^ of March and 2^nd^ of April) and both the pipelines, C1 and C2, using the newly designed primer sets specific for SARS-CoV-2. The analyzed sequences showed, for both ORF1ab and S partial gene regions, 100% identity with the first SARS-CoV-2 sequence detected in Italy (MT066156), isolated on 30^th^ January 2020 from a Chinese tourist by the Institute “Lazzaro Spallanzani” (INMI, Rome). Given the high level of conservation of the two analysed regions, 100% identity was also detected with several sequences in GenBank and with all the other Italian SARS-CoV-2 genomes deposited in Gisaid.

Significantly, on February 24^th^ and 28^th^, when the samples positive for SARS-CoV-2 were collected in Milan, COVID-19 infections were still limited in Italy, the first Italian autochthonous SARS CoV-2 positive case having been reported only a few days earlier, on February 21^st^. On February 28^th^, the total number of SARS-CoV-2 positive patients reported in all Italy was only 888,with 531 (57%) in Lombardy, the most affected region in the country. However, at that time, the vast majority of cases in Lombardy were recorded in the provinces of Lodi, Cremona and Bergamo (182, 123, and 103 cases, respectively). In comparison, in the province of Milan (an even larger area compared to the metropolitan area served by the selected WWTPs) only 29 cases had been reported (Dipartimento della Protezione Civile, Bulletin 28.02.2020).

These results provide evidence of the sensitivity of environmental surveillance for the detection of ongoing outbreaks in the population. Virus detection in sewage, despite the low incidence of reported human infections, may be associated with the ability of environmental surveillance to detect mild, subclinical, or asymptomatic cases. These infected individuals shed viruses into local sewage systems and contribute to virus circulation while remaining substantially undetectable by clinical surveillance, a phenomenon known as the “surveillance pyramid” (Martinez Wassaf et al., 2014). Clinical surveillance, indeed, only captures the tip of the iceberg of viral diseases (hospitalized patients or laboratory diagnosed cases). In contrast, monitoring of urban wastewaters makes it possible to capture the full extent of the diseases at a community level.

As regards to the influent samples collected in Rome, SARS-CoV-2 detection was obtained on March 31, when the epidemic had spread considerably in Italy. In that date, a total of 77.635 SARS-CoV-2 infections had been reported in Italy, of which 3.095 in Latium Region and 2.186 in the province of Rome (Dipartimento della Protezione Civile, Bulletin 31.03.2020), with about 85% of them being active cases (Assessorato alla Sanità e all’Integrazione Socio Sanitaria della Regione Lazio; Ministero della Salute, Bulletin 31.03.2020). Given this diffusion of the virus, with such several excreting patients (symptomatic and asymptomatic), the detection of the viral RNA in the tested samples is not surprising and, consistently, the samples taken two days later, on the 2^nd^ April, at the same WWTP were still positive for SARS-CoV-2 RNA.

Following this investigation on the occurrence of SARS-CoV-2 RNA in sewage, the production of quantitative data on virus concentration in raw sewage will be undertaken. This approach will allow obtaining a rough estimation of the total number of subjects excreting the virus, by integrating - as done by Wu and co-workers in samples taken in the United States (Wu et al., 2020) - the available information on viral shedding rates, WWTPs loads, and virus concentration in wastewaters. Moreover, the environmental surveillance will be extended to the collection of wastewater samples available in the Department of Environment and Health of the Italian National Health Institute, that were collected throughout Italy in the framework of different projects on enteric viruses. Such monitoring will provide a picture of the SARS-CoV-2 circulation across the different regions of Italy and over time, to better understand the virus circulation, as provided by wastewater-based epidemiology (WBE) and compare it to the clinical data. Samples collected before the reporting of the first known Italian case on February 21 will also be tested, to possibly infer when SARS-CoV-2 first appeared in Italy. In a previous study, indeed, wastewater monitoring provided evidence that a novel variant of Norovirus GII.17 (termed Kawasaki 2014) had been circulating in the Italian population before its first appearance and identification in clinical cases, later becoming one of the prevalent variants in the population (Suffredini et al., 2018).

Also and most important, environmental monitoring of SARS-CoV-2 in sewages will continue when the emergency phase is over, and its circulation in the population will be considered limited. Indeed, sewage surveillance could also serve for the early detection of a possible re-emergence of COVID-19 in urban areas. WHO recommends environmental surveillance for poliovirus as an early warning system. As an example, during 2013, Israel observed the silent reintroduction and transmission of wild poliovirus type 1, detected through routine environmental surveillance performed on sewage samples (https://www.who.int/csr/don/2013_09_20_polio/en/) without the reporting of any clinical cases. Environmental monitoring, therefore, appears to be an effective measure for proving early warning against pathogen reintroduction.

In conclusions, the main findings of this study are:

1. first detection of SARS-CoV-2 RNA fragments in sewage in Italy;
2. demonstration of the suitability of the WHO protocol for sewage treatment to enveloped viruses after appropriate modifications;
3. design of a novel nested PCR assay specific for SARS-CoV-2, useful for screening purposes.

Further research will clarify the applicability of WBE to SARS-CoV-2 for prompt detection, the study, and the assessment of viral outbreaks.

## Data Availability

Data Availability

